# *PEStimate*: Predicting offspring disease risk after Polygenic Embryo Screening

**DOI:** 10.1101/2025.09.05.25335168

**Authors:** Liraz Klausner, Ateret Revital, Todd Lencz, Shai Carmi

## Abstract

**Motivation:** Polygenic embryo screening (PES) is a new technology in reproductive medicine, whereby human in-vitro fertilization (IVF) embryos are screened for their genetic risk of complex, polygenic diseases. PES aims to reduce the burden of polygenic diseases in offspring by prioritizing the selection of low-risk embryos. However, given that polygenic diseases are usually late-onset, PES outcomes cannot be evaluated empirically and must be estimated by epidemiological modeling. The commonly used liability threshold model has been previously used to predict PES outcomes. However, predictions rely on complex sets of equations, some of which require numerical integration or simulation. Further, previous models failed to account for the possibility that the selected embryo will not be born.

**Results:** Here, we present *PEStimate*, a freely available online app for predicting PES outcomes when screening for a single disease. *PEStimate* predicts the offspring risk with and without PES, as well as plots of the risk reduction vs key parameters. Users can adjust the number of available embryos, the live birth rate, the disease prevalence and heritability, the accuracy of the genetic risk predictor, the embryo selection method, and the genetic risk and disease status of the parents and other relatives. Our new model for PES, which includes the possibility of embryo implantation failure, shows that risk reductions have been previously overestimated. *PEStimate* provides geneticists, healthcare professionals, patients, policymakers, and other stakeholders a necessary tool for examining the impact of PES and weighing its potential benefits against expected personal and societal harms.

**Availability and Implementation:** *PEStimate* is freely available at: https://polygenicembryo.shinyapps.io/pestimate. The source code is available at: https://github.com/Lirazk/PEStimate.

## Introduction

Evaluation of the genetic composition of in-vitro fertilization (IVF) embryos, called preimplantation genetic testing, has been used for several decades to avoid pregnancies with embryos carrying pathogenic variants or chromosomal aberrations^1^. Developments in whole-genome amplification, sequencing, and genotyping methods have made it feasible and affordable to generate genome-wide data for embryos^2–9^. Recently, multiple companies started offering IVF patients screening of their embryos for complex, polygenic diseases, including, for example, coronary artery disease, hypertension, diabetes (types 1 and 2), breast and prostate cancers, inflammatory bowel disease, and schizophrenia^10–15^. Each of these conditions is influenced by thousands of genes and variants, in addition to non-genetic risk factors. The genetic risk for each condition is usually estimated as a polygenic risk score (PRS), a weighted count of the number of risk-associated alleles carried by an individual^16,17^. Embryos are then prioritized for transfer to the uterus of the IVF patient by their PRS for one or more conditions.

Polygenic embryo screening (PES) has been heavily criticized by multiple professional societies and authors for possible harms to patients and society. Potential harms to patients include discarding viable embryos, an unnecessary exposure to IVF, insufficient counselling, choice overload, and possible long-term adverse psychological impact. Ethical and social harms include a slippery slope towards eugenics and selection for traits, increasing stigmatization and discrimination, unequal access and benefits across populations, distraction from non-genetic causes of disease, commercialization and overmedicalization of reproduction, and changing reproductive norms and parent-child relationships^18–27^.

Despite these concerns, PES may reduce disease burden in the next generation. Given that most of the diseases screened develop in adolescence or adulthood, risk reduction cannot be determined experimentally, but must be predicted based on epidemiological models. A number of studies used the liability threshold model (see Methods) to predict the risk reduction that could be achieved with PES^14,15,24,28^. The results suggest that PES may lead to significant risk reductions, at least when having many embryos and under ideal conditions, of up to 50% when screening common diseases such as schizophrenia or Crohn’s disease. Risk reduction can also be estimated by comparing disease outcomes between adult siblings, by selecting between the siblings as if they were IVF embryos. An analysis of sibling pairs from the UK biobank predicted large risk reductions even when siblings are prioritized based on their combined genetic risk for multiple diseases^11,29^.

To facilitate informed decision-making by patients, clinicians, researchers, and policymakers, it is crucial that all stakeholders have access to clear and evidence-based information on PES. However, the model-based predicted risk reductions rely on a series of complex equations that require numerical multiple integration for each setting of interest. Alternatively, risk reductions based on biobank data are limited to the specific conditions documented in the biobank, the currently existing PRS, and a comparison of usually only just two siblings.

Risk reduction calculators are currently available only from companies offering PES: Genomic Prediction^30^, Orchid Health^31,32^, and Herasight^15,33^. These calculators leave multiple gaps. The Genomic Prediction calculator is based on *unrelated* individuals from the UK Biobank. However, given that PRS is more variable among unrelated individuals than it is among sibling embryos, risk reductions are overestimated^29^. Further, the calculator is limited to specific phenotypes, scores, and UK-based European-ancestry individuals. The calculator by Orchid is based on pre-computed simulations of the liability threshold model, and it is thus again limited to specific phenotypes and scores. The Orchid calculator permits specification of the disease status of some of the first- and second-degree family members of the embryo. The Herasight calculator is also limited to pre-computed model simulations of specific phenotypes. It permits specification of parental disease history and ancestry, as well as and ancestry- and sex-specific disease prevalence and heritability.

In addition to the above-mentioned gaps, all currently available risk reduction estimates assume that an embryo will be born once transferred. However, even chromosomally intact embryos will often fail to implant or will result in a miscarriage^34^. Thus, it is the number of potential *births* per IVF cycle, rather than the number of *embryos*, which determines the predicted risk reduction. However, the number of births will not be known to IVF patients at the stage of embryo screening. Therefore, more realistic models for PES must take into account the uncertain nature of embryo implantation outcomes.

Here, we present *PEStimate*, an online app for estimating risk reduction for embryo selection for a single disease. *PEStimate* predicts risk reductions based on analytical solutions to the liability threshold model^28^, and it is therefore fast and flexible. The online interface permits users to estimate the risk reduction for any given parameter set, as well as plot graphs of the risk reduction vs key parameters. Further, we incorporate, for the first time, embryo birth parameters into the PES model, demonstrating that risk reductions were previously overestimated. A preliminary version of the app has been mentioned, without additional information, as part of our 2024 review paper^18^. Here, we present a much-expanded version of the app, along with complete details on its underlying models, implementation, and user interface. We believe that *PEStimate* will become a valuable tool for researchers, clinicians, patients, and other stakeholders who are interested in evaluating the prospects of polygenic embryo screening.

## Methods

The Supplementary Material provides complete details on our modeling assumptions and the derivation of the risk reduction under each setting. Here, we briefly describe the key components of our models.

### The setting

We assume the availability of *n* embryos, generated by IVF patients in a single oocyte-retrieval cycle, who have all developed to the blastocyst stage. The genome of each embryo is sequenced or genotyped, and the embryos are all euploid (i.e., free of chromosomal aberrations). The embryos are then screened for a prespecified disease with prevalence/lifetime risk *K*. The disease is assumed to be polygenic, with a normally distributed PRS explaining a proportion *r*^2^ of the variance in disease liability (see below) in the target population. The target population may be of any ancestry, including non-European. We assume that *K* and *r*^2^ are known from published disease-specific studies. It is possible to convert^15,35,36^ multiple metrics of PRS accuracy (e.g., the AUC) into *r*^2^. The value used for *r*^2^ should represent only the impact of direct, within-family effects, i.e., not incorporating confounding due to, e.g., population structure, assortative mating, or correlation with parental genotypes^37^.

### The genetic risk model

We use the liability threshold model^38,39^, a popular framework for modeling and predicting risk in genetic epidemiology^40–46^. The model assumes that the binary disease status is determined by an underlying, unobserved, continuous liability, and that individuals with liability above a threshold are affected. In the context of embryo selection, the model can be written, for *n* embryos^28^, as

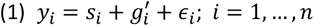

In Eq. (1), *y*_*i*_ is the (total) liability of embryo *i* for the specific disease screened. Assuming that the liability is distributed as a standard normal variable, and given the disease prevalence *K*, the disease threshold is *z*_*K*_ = Φ^−1^(1 − *K*), where Φ(·) is the cumulative distribution function (CDF) of the standard normal distribution.

The liability in Eq. (1) is a sum of genetic and non-genetic risk factors. *s*_*i*_ is the PRS (or “score”) of embryo *i*; 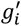 represents any remaining genetic factors not captured by the score; and *ϵ*_*i*_ represents non-genetic, random, or environmental risk factors. All components are assumed to be independent and normally distributed with zero means. The variance of *s*_*i*_ is denoted as *r*^2^, which is also the proportion of variance in liability explained by the PRS. It is used as a measure of the PRS accuracy. The variance of 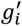 is *h*^2^ − *r*^2^, such that the overall variance of the genetic risk factors is equal to the heritability *h*^2^. The variance of *ϵ*_*i*_ is the remaining 1 − *h*^2^.

We assume that non-genetic risk factors are independent between family members, and similarly for the genetic risk factors of the parents. In contrast, all genetic factors have correlation 1/2 between first degree genetic relatives (e.g., pairs of embryos or an embryo and a parent). This results in the following form of the liability of each embryo^28^,

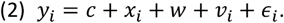

In Eq. (2), the PRS is broken down into *s*_*i*_ = *c* + *x*_*i*_, where *c* is *shared* between all embryos, while *x*_*i*_ ∼ *N*(0, *r*^2^/2) is an embryo-specific component. It can be shown^28^ that *c* = (*s*_*m*_ + *s*_*f*_)/2, where *s*_*m*_ and *s*_*f*_ are the maternal and paternal scores, respectively, i.e., *c* is the mean parental PRS. Similarly, 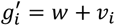, where 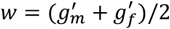 is the mean parental non-PRS genetic component and *v*_*i*_ ∼ *N*(0, (*h*^2^ − *r*^2^)/2) is embryo specific. Eq. (2) also holds for any previously born sibling of the embryos.

### Family information

The baseline model for the risk of the selected embryo assumes that the parents are random individuals from the population. However, it is also possible to specify the PRS of the parents and/or the disease status of either one or both parents and any number of siblings. The distribution of the genetic risk factors of the family members is then conditioned upon their disease status.

### Embryo selection

We consider two embryo selection strategies^28^. (i) *Lowest risk prioritization*: selection of the embryo with the lowest PRS (equivalently the lowest *x*_*i*_, for *i* = 1, …, *n*). (ii) High-risk exclusion: exclusion of all embryos with PRS above a cutoff (based on a PRS quantile *q*). If all embryos are high risk, a random embryo is selected. It is assumed that the selected embryo is transferred to the uterus of the patient for the purpose of initiating a pregnancy.

### The number of embryos/births

If an embryo is transferred but not born, its PRS is irrelevant for embryo screening. Therefore, the only parameter relevant for risk reduction in a given IVF cycle is the number of births. We derive the risk of the selected embryo under the following three models.

i. There are *n* embryos, and each embryo transferred is born. This model is unrealistic, but it provides a useful baseline. In this model, the number of births is fixed at *n*.
ii. There are *n* embryos, and each embryo transferred is born with probability *p*. In this model, the number of births is binomial with parameters (*n, p*).
iii. The number of embryos is Poisson distributed with mean *n*, and each embryo transferred is born with probability *p*. In this model, the number of births is Poisson with mean *np* (see the Supplementary Material, Section 5).

For models (ii) and (iii), we assume that at least one birth was achieved, or otherwise the risk reduction is undefined. Equivalently, if the IVF cycle resulted in no live births, it is assumed that additional cycles were performed until at least one birth was achieved. These models are implemented in the app only for the lowest risk prioritization strategy.

### Estimating the risk reductions

Our goal is to estimate the risk of the selected embryo, i.e., the probability of the embryo to be affected as an adult. Under each of the settings described above, we analytically derive the risk of the selected embryo in the Supplementary Material or use the published result^28^. The risk is always given as a single or a multiple integral. The Supplementary Material describes how *PEStimate* solves these integrals numerically or using a Monte-Carlo approach. We implemented all numerical methods in R (version 4.5.1) and the user interface in R Shiny (version 1.10.0).

We report the following risk reduction metrics. The absolute risk reduction is defined as

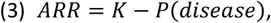

where *K* is the disease prevalence (i.e., the risk of a random individual/embryo from the population) and *P*(*disease*) is the risk of the selected embryo.

The relative risk reduction is defined as

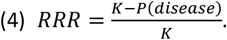

The number of patients that would need to be screened in order to avoid a single disease case is computed as *NNS* = 1/*ARR*.

## Results

### The impact of modeling the possibility of embryo transfer failure

All previous models for PES outcomes^14,15,24,28^ reported risk reductions vs the number of (euploid) embryos being screened, implicitly assuming that the transferred embryo will be born. However, given that many euploid embryo transfers fail to result in live birth, the availability of a given number of embryos does not guarantee the predicted risk reductions. Thus, the number of embryos must be replaced with the number of *potential births* from all embryos generated in the egg retrieval cycle.

However, for any given number of embryos, the number of births is not known prior to embryo transfer, and it is better modeled as a random variable.

A naïve approach to modeling risk reduction may be setting the number of births to the *expected (mean) number of births* given the embryo count or other parental parameters. However, in the Supplementary Material, we prove that

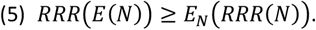

In Eq. (5), *N* is the number of births and *RRR* is the relative risk reduction (see Methods; Eq. (4)). The inequality demonstrates that using the mean number of births, *E*(*N*), and then predicting the corresponding relative risk reduction (i.e., *RRR*(*E*(*N*))) always overestimates the risk reduction compared to properly averaging the risk reduction over all possible numbers of births (*E*_*N*_(*RRR*(*N*)). Thus, the naïve approach would lead to over-optimistic predictions.

To address this problem, we propose two new models for the number of births. In the first, the number of embryos is given, and each embryo is born with a certain live birth probability (the “binomial” model; see Methods). In the second, the number of embryos is also random (the “Poisson” model).

We compare predictions from the three models – the binomial model, the Poisson model, and fixing the number of births to its mean – in Figure 1. To allow a fair comparison, we selected model parameters in a way that the mean number of births is the same across models (see the Supplementary Material). As predicted by Eq. (5), using a fixed number of births indeed overestimates the risk reduction. Overestimation is substantial up to ≈4-5 average births per cycle. The predictions of the two models with a random number of births are numerically similar. These observations are consistent across various values for the prevalence and the PRS accuracy.

**Figure 1.**
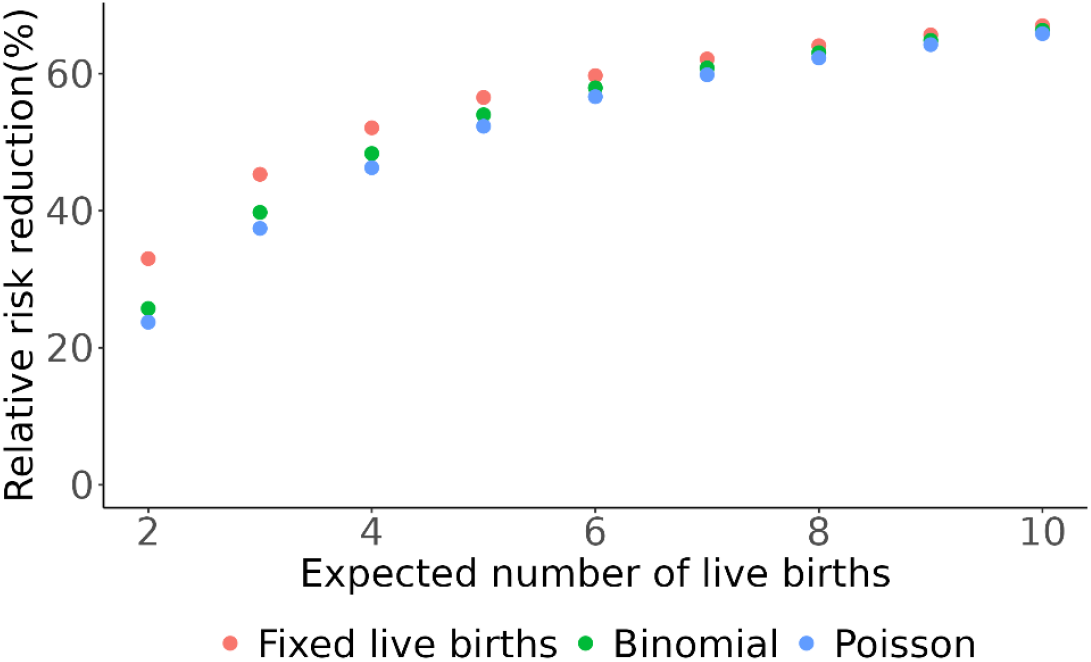
A comparison of relative risk reductions predicted by three models for the number of births. We consider the lowest risk prioritization selection strategy and assume at least one birth. We set the live birth rate to *p* = 0.4, the prevalence to *K* = 1%, and the PRS accuracy to *r*^2^ = 10%. We set the other model parameters such that the mean number of births is the same across all three models (see the Supplementary Material).

### An online risk reduction estimation app

We implemented an online graphical user interface for the estimation of risk reductions as an R Shiny app. The app has three pages.

The first is an “About” page, providing a brief description of the app, the embryo selection strategies, the input and output parameters, references, and contact information.

The two other pages are “Plot” and “Calculator”. On the first visit to each of those pages, the user is presented a popup window with “Important notes”. Most of these notes inform the user on the model’s assumptions, as described in the previous sections. The last note indicates that polygenic embryo screening has been associated with ethical, social, and legal problems. The user must confirm reading and understanding the notes, and that the app is intended for research purposes only.

The “Calculator” page (Figure 2) allows users to input all relevant parameters and examine the risk reduction results. Most input parameters have an accompanying tooltip. The input parameters are organized in four panels.

**Figure 2.**
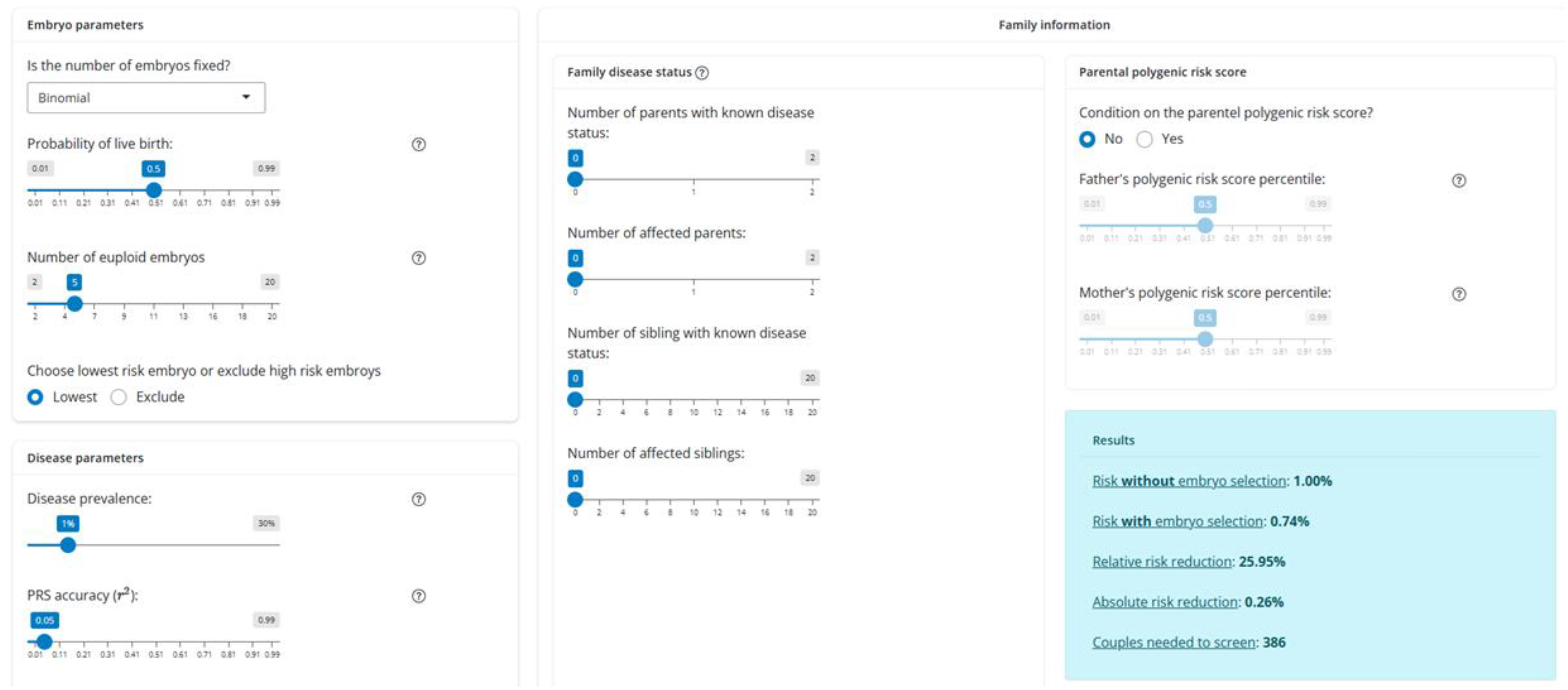
A screenshot of *PEStimate*’s Calculator page. The “Plot” page (Figure 3) allows users to plot the relative and absolute risk reductions vs some of the model’s parameters. The model for the number of embryos/births is specified as in the Calculator page. Users can then plot the risk reduction against each of these parameters: (i) The number of births/embryos (depending on the model). (ii) The disease prevalence. (iii) The PRS accuracy (*r*^2^). (iv) For the high-risk exclusion strategy, the exclusion cutoff (as a PRS percentile). To limit the complexity of the Plot page, it does not permit specification of family information.

The “Embryo parameters” panel first requires users to select a model for the number of embryos/births from among the three models described in the Methods section. For a fixed number of births, the user needs to specify the number. For the binomial number of births, the user needs to specify the number of (euploid) embryos and the birth rate per embryo transfer. For the Poisson model, the user needs to specify the *mean* number of embryos and the birth rate. Finally, the user needs to specify the embryo selection strategy (lowest risk prioritization or high-risk exclusion; see Methods).

The “Disease parameters” panel specifies the disease prevalence (or the probability of a random embryo to be affected) and the accuracy of the PRS (*r*^2^, the proportion of variance in liability explained by the PRS; see Methods). If the high-risk exclusion strategy is selected, the user must also specify a PRS percentile cutoff. If the disease status of family members is provided (see next), the user must specify the heritability *h*^2^ (the proportion of variance in liability explained by genetic risk factors; see Methods; *h*^2^ must be greater than *r*^2^). These parameters must be obtained from the relevant literature or in-house data.

The final pair of panels provide information on family members. The “Family disease status” panel specifies the number of parents and already-born siblings (of the embryo) with a known disease status, and then the number of affected parents and siblings. The “Parental PRS” panel allows users to input the paternal and maternal PRS percentiles.

The output of the model (light blue box in Figure 2) provides the following information. (i) The risk of the future child *without* embryo screening. This is equal to the disease prevalence, except when family information is provided, in which case the risk is based on the given family parameters. (ii) The risk *with* embryo screening. (iii) The relative risk reduction; (iv) the absolute risk reduction; and (v) the number of patients that need to be screened in order to avoid a single disease case (see Methods). In case the user has provided the disease status of family members, the output (which is based on Monte Carlo sampling) also includes standard deviations.

### Examples

*Case 1*. One spouse in a couple is affected with schizophrenia and the other is unaffected. The couple is theoretically fertile but they have no children yet. What is the schizophrenia risk reduction that can be achieved for their future child by selecting the lowest risk embryo? We set the prevalence to *K* = 0.01^47^, the proportion of variance explained by the PRS to *r*^2^ = 0.07^48^, and the heritability to *h*^2^ = 0.8^49^. We assume that the couple will have a random (Poisson) number of euploid embryos with mean 4^50,51^ and we set the birth rate per euploid embryo transfer to 0.5^34,52^. The risk of the future child decreases from 8.33% without PES to 6.75% with PES.

*Case 2*. A couple already has one child who is affected with type 1 diabetes. The parents are healthy and fertile. The parents were tested, and their paternal and maternal PRS percentiles were at the top 5^th^ and 15^th^, respectively. They already went through IVF and generated five euploid embryos. What is the type 1 diabetes risk reduction that can be achieved for their future child by selecting the lowest risk embryo? We set the prevalence to *K* = 0.0015^53^, the PRS accuracy to *r*^2^ = 0.25^35,54^, the heritability to *h*^2^ = 0.83^55^, and the birth rate to 0.5. The risk of the future child deceases from 3.16% without PES to 1.58% with PES.

*Case 3*. A couple suffering from infertility went through IVF and generated a number of embryos that would lead to two live births. They have no children yet. They are interested in guaranteeing that their future child is not at high risk for Alzheimer’s disease (for which their own disease status is yet unknown), and thus they would like to exclude all embryos at the top 10% of genetic risk. We set the prevalence to *K* = 0.11^15^ and the PRS accuracy to *r*^2^ = 0.16^15^. In this setting, the risk of the future child deceases from 11% without PES to 10.0% with PES.

*Case 4*. A healthy, theoretically fertile, but yet childless couple is planning to go through IVF to reduce their future child’s risk for type 2 diabetes. They were not genetically tested and their disease status is yet unknown. They do not know how many euploid embryos they would need to have to achieve a given risk reduction. We set the prevalence to *K* = 0.20^15^, the PRS accuracy to *r*^2^ = 0.21, and the birth rate to 0.5. The risk reduction (relative and absolute) is shown in Figure 3 as a function of the number of embryos.

**Figure 3.**
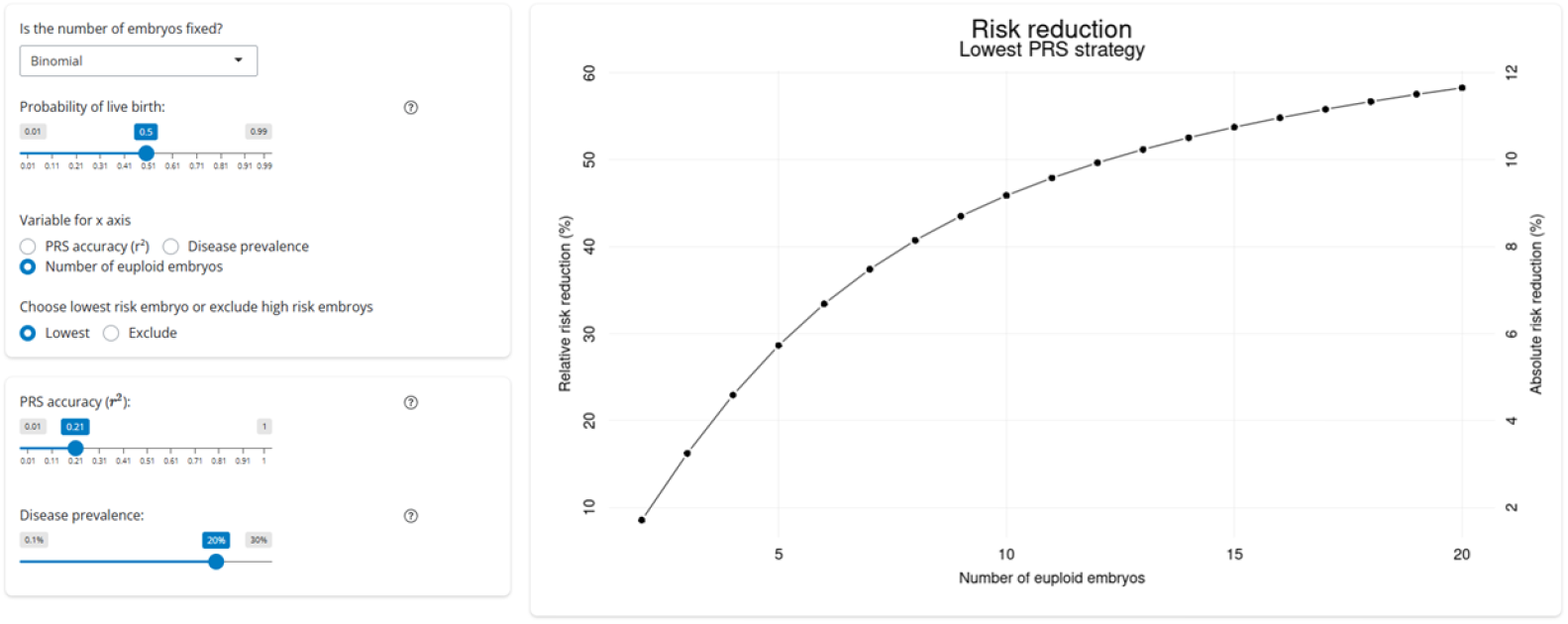
A screenshot of *PEStimate*’s Plot page. The parameters correspond to Case 4 in the Examples subsection.

## Discussion

PES is an emerging assisted reproductive technology with a rapidly evolving landscape and massive public interest^56–58^. It adds a highly contentious dimension to reproductive decision-making, raising significant ethical, clinical, and social concerns. Further, attitudes towards PES diverge between healthcare professionals, who are reluctant to offer the screen due to perceived low clinical utility and potential for harm, and members of the general public, who show both approval of PES and self-interest^18,59–61^. In this context, it is crucial to provide all stakeholders with an accessible tool for evaluating the potential utility of PES. *PEStimate* addresses this need by allowing users to examine PES outcomes under a wide array of parameters and models.

There are many limitations to our modeling framework, most of them previously discussed^28^. Briefly, the model relies on the assumption that disease status is truly binary (affected/unaffected), that each disease has an underlying continuous liability, and that only individuals with liability above a threshold are affected^62^. Further, it assumes a normal distribution of the liability components, a correlation of 0.5 between the genetic liabilities of first-degree relatives, and independence between the non-genetic risk factors of family members and between genetic and non-genetic risk factors. Therefore, results from our model should be considered a crude approximation. Nevertheless, the model’s predictions agree closely with simulations^28^, and our model and app provide crucial information regarding the impact of the various parameters.

Our approach for modeling a non-deterministic number of births is novel and avoids one of the main pitfalls of existing risk reduction estimators. We use our new model to show that previous risk reduction estimates were inflated, even after accounting for the possibility of transfer failure by using the expected number of births. Naturally, our model also relies on several assumptions. These include a constant live birth rate across families and across all embryos in a given IVF cycle, independence between the birth rate and other parameters, and that at least one birth was achieved. The model with a random number of embryos is limited to a Poisson distribution. However, our analytical framework (Supplementary Material Section 5) can seamlessly incorporate any other distribution of the number of live births. Extending the model and app to additional distributions (e.g., a negative binomial number of embryos) is thus straightforward.

The main limitation of our app is its focus on embryo selection for a single disease risk. Selecting embryos simultaneously based on their risk for multiple diseases introduces multiple complications, such as the need to model the genetic correlation between diseases and the explosion in the number of possible selection strategies. While the gain under simultaneous selection for multiple traits can be derived analytically^63^, no analogous result exists for disease risk, and estimating the risk reduction will require the development of a fast simulation approach. While we leave modeling of selection against multiple diseases for future work, biobank sibling data may provide a more attainable alternative for estimating risk reductions in the near future.

The main advantage of our app is the ability to perform a fast exploration of the parameter space. This is facilitated by the analytical solutions we derived under most settings, in contrast to existing calculators that rely on pre-computed simulations. One useful application is examining how PES outcomes are affected by reduced PRS accuracy, e.g., due to non-European ancestry, environmental or generational differences, or confounding by indirect effects.

In conclusion, *PEStimate* allows users to research what approximate risk reductions are expected from PES under various contexts, including different diseases, PRS, embryo counts, and family history. We believe our app will be useful for researchers, healthcare professionals, regulators, and the general public when considering the benefits and harms of the technology.

## Data Availability

All data produced in the present work are contained in the manuscript

https://polygenicembryo.shinyapps.io/pestimate/

## Glossary

*K*: The disease prevalence in the population.
Φ: The cumulative distribution function of the standard normal variable.
*ϵ*: The non-genetic component of the liability.
*ϕ*: The probability density function of the standard normal variable.
*arr*: The absolute risk reduction.
*c*: The PRS-specific genetic component of the liability shared between all embryos.
*g*: The (additive) genetic component of the liability (with variance *h*^2^).
*g*^*′*^: The non-PRS genetic component of the liability (with variance *h*^2^ − *r*^2^).
*h*^2^: The proportion of the variance in the liability due to all additive genetic factors; the *heritability*.
*n*: The number of embryos available for selection.
*nns*: The number of patients that would need to be screened to avoid a single case.
*r*^2^: The proportion of the variance in liability explained by the PRS (the PRS accuracy).
*rrr*: The relative risk reduction.
*s*: The PRS-specific genetic component of the liability (with variance *r*^2^).
*v*_*i*_: The non-PRS genetic component of the liability unique to embryo *i*.
*w*: The non-PRS genetic component of the liability shared between all embryos.
*x*_*i*_: The PRS-specific genetic component of the liability unique to embryo *i*.
*z*_*K*_: The upper quantile of the standard normal distribution corresponding to probability *K*.
*z*_*q*_: The upper quantile of the standard normal distribution corresponding to probability *q*.

## Acknowledgements

We thank Antonio Capalbo for Discussions.

## Author contributions

Conceptualization: SC; Formal Analysis: LK; Funding Acquisition: SC, TL; Investigation: LK, SC, AR, TL; Methodology: LK; Project Administration: SC; Software: LK; Supervision: SC; Visualization: LK, AR; Writing – Original Draft Preparation: SC, LK; Writing – Review & Editing: LK, SC, TL.

## Data and code availability

*PEStimate* is available at: https://polygenicembryo.shinyapps.io/pestimate. The source code is available at: https://github.com/Lirazk/PEStimate.

## Supplementary Material

## 1 Overview

This document provides technical details on the implementation of the polygenic embryo screening (PES) risk reduction calculator. The document is organized as follows. Section 2 presents a mathematical model for PES for a single disease [from our previous work (Lencz et al., 2021)]. The model defines the genetic and non-genetic risk factors of the embryo, how they are distributed across the embryos, and how they translate to disease risk. Section 3 describes the integrals that we numerically solve to provide the risk reduction for two selection strategies: lowest risk prioritization and high-risk exclusion. Section 4 describes a Monte Carlo method for estimating the risk reduction when the disease status of family members is known. Section 5 demonstrates how risk reduction can be computed even if the selected embryo is not guaranteed to be born and when the number of embryos is not known in advance.

## 2 A model for polygenic embryo screening

Our model is based on Lencz et al., 2021. We assume the availability of *n* embryos. It is assumed that all embryos are viable, have developed to the blastocyst stage around day-5 post-fertilization, and are euploid (as assessed using, e.g., PGT-A). In this section, we further assume that once the selected embryo is transferred to the uterus of the patient, it will lead to pregnancy and live birth. We relax this assumption in Section 5. We consider embryo selection based on the risk of a single disease.

We model the risk to the embryo using the liability threshold model (Falconer, 1965), under which it is assumed that a disease has an underlying, unobserved, continuous liability, and that the disease emerges if the liability exceeds a threshold. The liability is assumed to have a standard normal distribution.

We write the liability most generally as

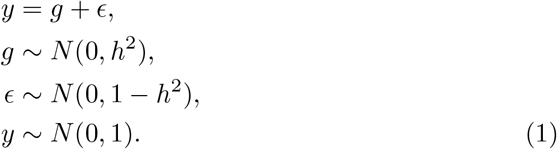

In Eq. (1), *g* is the additive genetic component of the liability and *ϵ* is the non-genetic component, which includes the environment and any random contributions. Both are assumed to be independent and normally distributed. The variance of *g*, or *h*^2^, is called the heritaiblity.

The genetic component *g* is usually unknown. We therefore write

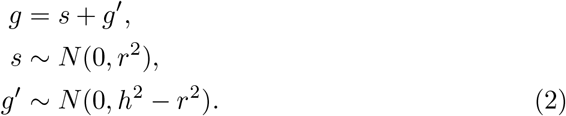

In Eq. (2), *s* is the polygenic risk score (PRS), which is computed using variants known to be associated with the disease based on the results of genome-wide association studies. *g*^*′*^ is the non-PRS genetic component, assumed to be independent of *s*. The variance of the PRS is *r*^2^, which is also the proportion of the variance of the liability explained by the PRS, and thus a measure of the PRS accuracy.

Therefore, for each individual,

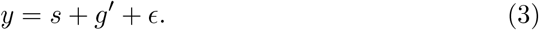

For a batch of *n* embryos, we split the contribution of each genetic component into shared and embryo-specific components. Following Lencz et al., 2021, we write the liability of embryo *i, y*_*i*_, as a sum of the following independent normal variables,

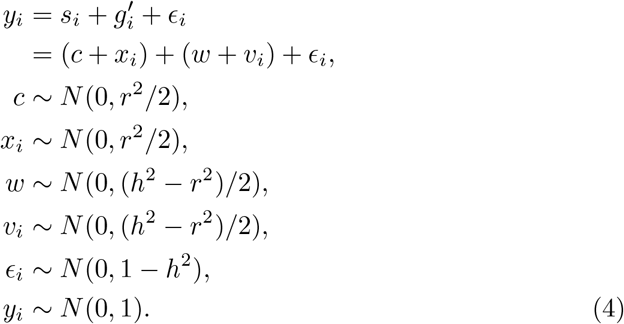

In Eq. 4, the PRS of embryo *i, s*_*i*_, is written as *s*_*i*_ = *c* + *x*_*i*_. *c* is the PRS-specific shared genetic component, which is identical between all embryos. It has variance *r*^2^*/*2, half the variance of the PRS in the population, which is a result of the correlation of 0.5 between any genetic value of full siblings (Wray et al., 2019). *x*_*i*_ is the embryo-specific PRS component, whose variance also equals to *r*^2^*/*2. Together, *s*_*i*_ = *c* + *x*_*i*_ is normally distributed with variance *r*^2^.

Similarly, the non-PRS genetic component of embryo *i*, 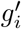, is written as 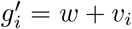, where *w* is the shared non-PRS genetic component, with variance (*h*^2^ −*r*^2^)*/*2, and *v*_*i*_ is the embryo-specific non-PRS genetic component (with the same variance). Together, 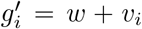 is normally distributed with variance *h*^2^ − *r*^2^. Finally, *ϵ*_*i*_ is the embryo-specific non-genetic component, and the total liability *y*_*i*_ is a standard normal variable. Note that all components are independent.

It can be shown that the shared PRS component *c* is equal to the mean of the maternal and paternal PRSs, *s*_*m*_ and *s*_*f*_, respectively,

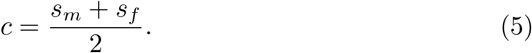

Similarly, the shared non-PRS component *w* is equal to the mean of the maternal and paternal non-PRS genetic components,

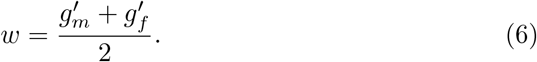

An individual is affected whenever its liability is above the disease threshold. To determine the threshold, we require

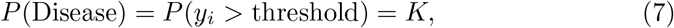

where *K* is the prevalence of the disease in the population. Given that *y*_*i*_ ∼ *N* (0, 1), the threshold is *z*_*K*_= Φ^−1^(1 − *K*), where Φ(*·*) is the cumulative distribution function of the standard normal variable.

We consider two embryo selection strategies (Lencz et al., 2021).

1. Lowest risk prioritization: Select the embryo *j* with the minimum PRS for the prespecified disease i.e., *j* = arg min_*i*_ *s*_*i*_ = arg min_*i*_(*c* + *x*_*i*_) = arg min_*i*_ *x*_*i*_.
2. High-risk exclusion: We exclude embryos if their PRS for the given disease is above a cutoff, *s*_*i*_ = *c* + *x*_*i*_ > *z*_*q*_*r*, where *z*_*q*_ is the PRS upper *q*-quantile, *z*_*q*_ = Φ^−1^(1 − *q*) (multiplied by *r* to generate the actual PRS cutoff). If one or more embryos remain (i.e., *s*_*i*_ < *z*_*q*_*r*), one embryo among them is selected at random. If no embryo has PRS below the cutoff, one embryo is selected at random from the entire set of *n* embryos.

Denote by *P* (Disease) the risk of the selected embryo, i.e., its probability to be affected with the disease as an adult. We consider a number of risk reduction metrics. The absolute risk reduction, *arr*, is defined as

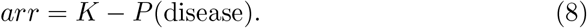

The relative risk reduction, *rrr*, is defined as

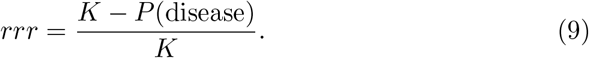

Finally, the number needed to screen, *nns*, is the number of IVF patients that would have to screen their embryos to avoid one future disease case. It is computed as *nns* = 1*/arr*.

See the Glossary for all parameter definitions.

## 3 Direct integration

As we showed in Lencz et al., 2021, the risk reduction across settings can be expressed as a multiple integral. In all settings except those when disease status is known for any family member, we solve these integrals numerically without resorting to simulations. We describe these integrals here. In Section 4, we address scenarios with a known family disease status.

### 3.1 Lowest risk prioritization

The disease risk when transferring the embryo with the lowest risk for the given disease is given by Eq. (20) from the appendix of Lencz et al., 2021,

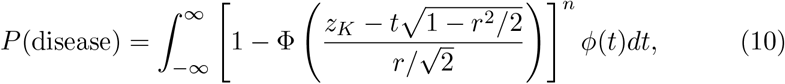

where *ϕ*(*·*) is the probability density function of the standard normal variable. The disease risk conditional on the mean parental score, *c*, is given by Eq. (23) in Lencz et al., 2021,

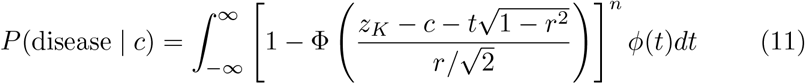

We solve these two integrals numerically using R’s `integrate` function, as in Lencz et al., 2021, as this approach is fast and sufficiently accurate.

### 3.2 High-risk exclusion

Given the mean parental score, the disease risk of the selected embryo when excluding high-risk embryos is given by Eq. (29) in Lencz et al., 2021. We also solve it using R’s `integrate`. The unconditional disease risk is given by Eq. (31) therein, which we solve as follows. First, we rewrite the (originally discontinuous) integral as

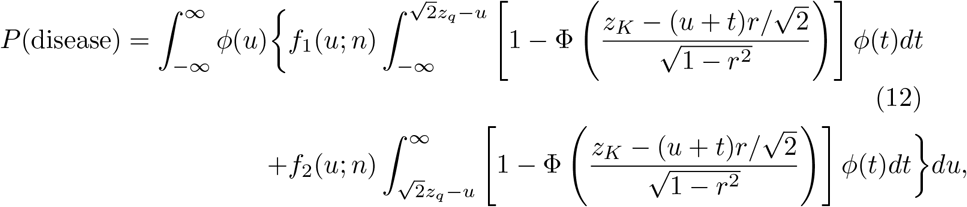

where *f*_1_(*u*; *n*) and *f*_2_(*u*; *n*) are defined as

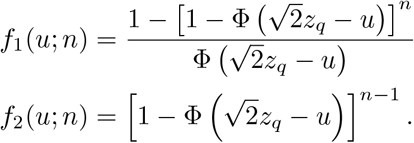

To proceed, we rewrite the inner integrals of Eq. (12) in a more generic form

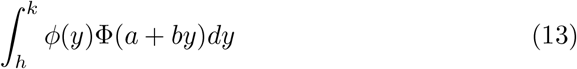

by using the identity 1 − Φ(− *y*) = Φ(*y*). This generic form of the integral is given in Owen, 1980. To solve the integral, we first show that it can be written in the following form,

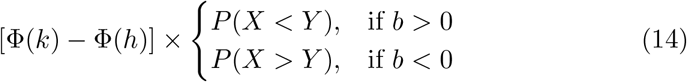

with

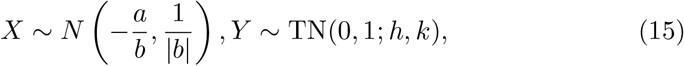

where TN(0, 1; *h, k*) denotes the truncated standard normal distribution with range (*h, k*), and *X* and *Y* are independent. To prove Eq. (14), we first consider *b* > 0,

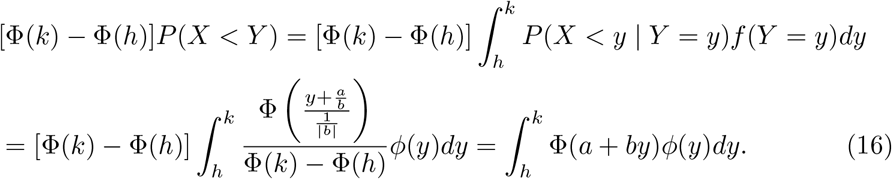

When *b* < 0, we instead calculate

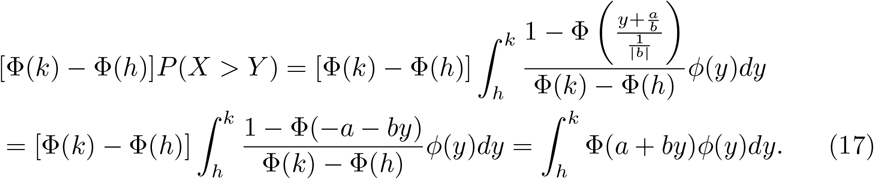

Given the structure of Eq. (12), we henceforth assume *b* > 0. To find the probabilities in Eq. (14), we write *Y* ^*′*^ ∼ *N* (0, 1) and *Z* = *X* − *Y* ^*′*^. The joint distribution of *Z* and *Y* ^*′*^ is

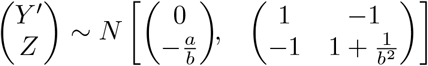

Back to the probability in Eq. (14), we need to calculate

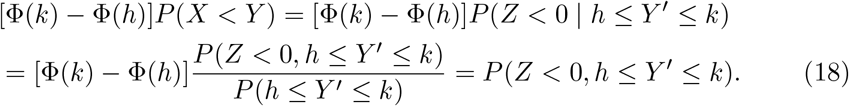

We thus only need the cumulative distribution function (CDF) of a bivariate normal variable, that is

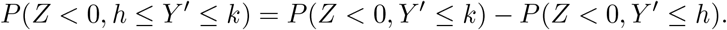

We next define a normalized 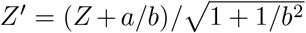, or 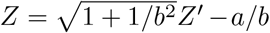. The correlation between *Z*^*′*^ and *Y* ^*′*^ is cor(*Z*^*′*^, *Y* ^*′*^) = cor(*Z, Y* ^*′*^) *≡* 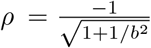. This gives

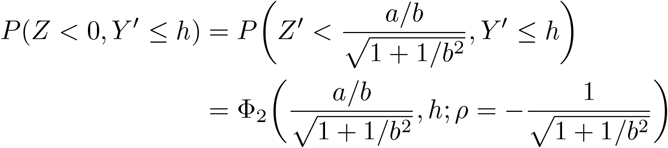

and similarly for *P* (*Z* < 0, *Y* ^*′*^ *≤ k*). Above, Φ_2_(*·,·*; *ρ*) is the CDF of a bivariate standard normal distribution with correlation *ρ*. This CDF can be calculated through Owen’s T function (Owen, 1956), which is defined as

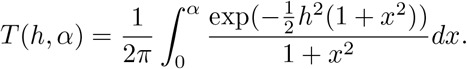

Fast and accurate methods exist for computing the T function numerically (OwenQ R package). Given the T function, the bivariate CDF can be computed as

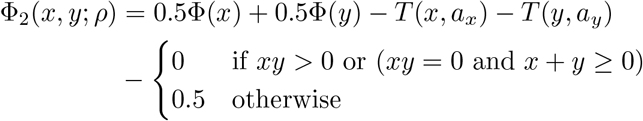

with

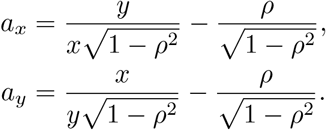

In Eq. (12), the inner integral was calculated using the T function, where some of the terms were simplified when we substituted *x* = −*∞* or *y* = *∞*. We computed the outer integral with R’s integrate function.

## 4 Conditioning on family history

The calculator can also incorporate information on the disease status of parents and siblings of the embryo. This is done by sampling the underlying genetic and environmental liability components of family members from their joint distribution, subject to linear constraints imposed by their disease status. We assume that the non-genetic risk factors are not shared between family members.

It is sometimes difficult to determine whether an individual is unaffected or *not yet* affected. Our model does not include the possibility of censoring. Thus, in settings involving late-onset diseases and relatively young family members, it is better not to condition on the disease status.

### 4.1 Setup

Let the vector *ℓ* contain all relevant random liability components for the family members considered. We limit our attention to parents and siblings of the embryo.

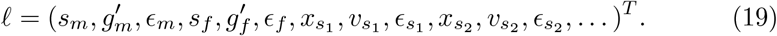

In Eq. (19), *s*_*j*_, 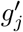, *x*_*j*_, *v*_*j*_, *ϵ*_*j*_ are the liability components, following the notation of Section 2, for person *j*,where *j* can be the mother (*m*), the father (*f*), or a sibling (*s*_1_, *s*_2_, …). We use the index *j* for the family members as the index *i* was used for the embryos.

Under the model of Section 2, *ℓ* follows a multivariate normal distribution, *ℓ* ∼ *N* (***µ*, Σ**). Before conditioning on the disease status of family members, the mean vector is ***µ*** = **0**. In the covariance matrix, **Σ**, the variance terms are *r*^2^, (*h*^2^ − *r*^2^), 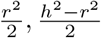, and 1 − *h*^2^ for each *s,g*^*′*^,*x,v*, and *ϵ*, respectively. The covariance terms are all zero (assuming no assortative mating and no shared environment), because for the siblings we only considered child-specific components.

### 4.2 Constraints due to known disease status of family members

The liability *y*_*j*_ of a family member *j* is a linear combination of the components of *ℓ*. Specifically, for the parents, *y* = *s* + *g*^*′*^ + *ϵ*. For the siblings, 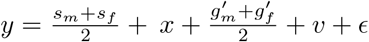. The disease status of family members imposes a constraint on their liability,

- If individual *j* is affected: *y*_*j*_ *≥ z*_*K*_.
- If individual *j* is unaffected: *y*_*j*_ < *z*_*K*_, which is equivalent to −*y*_*j*_ > −*z*_*K*_.

The constraints for all individuals with a known disease status can be combined into a matrix inequality **G***ℓ ≥* **r**, where each row of **G** defines the linear combination for one individual’s liability (negated for unaffected individuals), and **r** contains the corresponding thresholds (*z*_*K*_ or −*z*_*K*_). For example, if sibling 1 is affected, *y*_*s*_ > *z*_*K*_, which means that 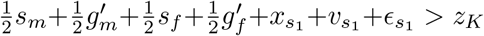, so that row of **G** would be (1*/*2, 1*/*2, 0, 1*/*2, 1*/*2, 0, 1, 1, 1, 0, …, 0). When the parental PRS is known, the known *s*_*m*_, *s*_*f*_ are used instead of being sampled.

**Figure S1.** Sampling *ℓ ∼ N* (*µ*, Σ) subject to G*ℓ ≥* r. **Input:** Mean ***µ***, Covariance **Σ**, Constraint matrix **G**, Constraint vector **r**. **Output:** A sample *ℓ* satisfying the constraints. 1. **Sample the constrained variables:** Draw a sample **y**^*′*^ = **G***ℓ ∼ N* (**G*µ*, GΣG**^*T*^) such that **y**^*′*^ *≥* **r**. *(Note: This step requires a specialized multivariate truncated normal sampler*.) **2. Sample an unconditional base:** Draw an independent sample **z** from the original, *unconditional* distribution: **z** *∼ N* (***µ*, Σ**). **3. Compute the conditional sample:** Adjust the unconditional sample **z** to match the sampled constrained value **y**^*′*^: *ℓ* = z + **ΣG**^*T*^ (G**ΣG**^*T*^)^−1^(y’ − **Gz**)

### 4.3 Sampling from the constrained distribution

We need to sample *ℓ* from its distribution conditional on **G***ℓ ≥* **r**. We use a Monte Carlo approach based on the algorithm of Cong et al., 2017. The algorithm is described in Figure 1. The justification relies on decomposing the sampling process: first sampling the constrained variable **y**^*′*^ = **G***ℓ* from its valid (truncated) range, and then sampling *ℓ* conditional on the drawn value **y**^*′*^. Sampling from multivariate truncated normal was implemented with the tmvtnorm package (Wilhelm and G, 2023). Other options for generating samples exist (Li and Ghosh, 2015, Cong et al., 2017).

### 4.4 Estimating the risk of the selected embryo

We run the algorithm of Figure 1 repeatedly to generate *K* independent samples, {*ℓ*^(1)^, …, *ℓ*^(*K*)^}, from the conditional distribution. For each sample *ℓ*^(*k*)^, we first define the parental components 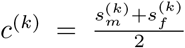 and 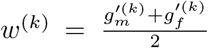. These define the shared components for potential embryos. Given *c*^(*k*)^ and *w*^(*k*)^, the liability of embryo *i* is 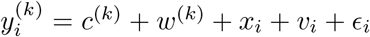. The embryo-specific components (*x*_*i*_, *v*_*i*_, *ϵ*_*i*_) are sampled independently from their respective distributions: *N* (0, *r*^2^*/*2), *N* (0, (*h*^2^ − *r*^2^)*/*2), and *N* (0, 1 − *h*^2^). We then simulate *n* such embryos. For the lowest-risk prioritization strategy, we select the embryo with the lowest *s*_*i*_. For the high-risk exclusion strategy, we randomly select an embryo after excluding embryos with PRS above the exclusion cutoff. We then determine whether the liability of the selected embryo is above the threshold (i.e., the embryo is affected).

To reduce the variance of the above method, and given that embryo selection depends only on the PRS *s*_*i*_, we only sample the embryo-specific PRS *x*_*i*_. We then analytically compute the risk of the selected embryo by integrating over *v*_*i*_ and *ϵ*_*i*_. Specifically, given that (for simulation *k*) *y*_*i*_ = *c* + *w* + *x*_*i*_ + *v*_*i*_ + *ϵ*_*i*_, we set 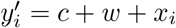 based on the simulated data, such that *y* = *y*^*′*^ + *v*_*i*_ + *e*_*i*_. Given that *v*_*i*_ and *ϵ*_*i*_ are independent, their sum has normal distribution *N* (0, (*h*^2^ − *r*^2^)*/*2 + 1 − *h*^2^) = *N* (0, 1 − *h*^2^*/*2 − *r*^2^*/*2). Thus, 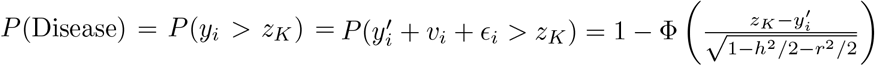.

## 5 Risk reduction with a variable number of embryos and births

### 5.1 Introduction

We have so far assumed that (*i*) the selected embryo is always born; and (*ii*) the number of embryos is given. Therefore, we computed the risk reduction as a function of the (fixed) number of embryos *n*. However, in practice, (*i*) only half or so of the embryos transferred are born (Cimadomo et al., 2023); and (*ii*) stakeholders may be interested in the *expected* risk reduction when the number of embryos is not known in advance (but rather only its distribution).

In this section, we derive the risk reduction for the case when either the number of embryos or the number of births (or both) are random variables. We only consider the lowest risk prioritization strategy; the case of the high-risk exclusion strategy is analogous but more tedious.

### 5.2 Why does it matter if the number of births is random?

Recall that the relative risk reduction is defined as *rrr* = (*K*− *P* (Disease))*/K* (Eq. (9)), where *K* is the disease prevalence and *P* (Disease) is the risk of the selected embryo (as computed in previous sections). More generally, denote the relative risk reduction as a function of the number of embryos as *rrr*(*n*). Under our initial assumptions, (*i*) *n* is the number of (live) births, because all embryos can be born; and (*ii*) *n* is a constant. In reality, these assumptions do not hold.

Naturally, an embryo transferred but not born is irrelevant for risk reduction. Therefore, the risk reduction is determined by the number of births. In the most general setting, the number of births is a random variable, denoted *N*. Naively, we could replace *rrr*(*n*), for a fixed *n*, with *rrr*(*E*(*N*)), where *E*(*N*) is the mean number of births.

However, we prove that in the case of the lowest risk prioritization strategy

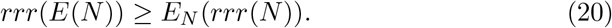

In Eq. (20), *E*_*N*_ (*rrr*(*N*)) is the mean risk reduction over all possible values of *N*. In other words, the risk reduction achieved by assuming that all IVF cycles result in the same number of births (the mean *E*(*N*)) is greater than the risk reduction when it is (more realistically) averaged over all possible values of *N*. Thus, not taking into account the uncertainty in the number of births overestimates the risk reduction.

To prove Eq. (20), it is sufficient to show that *rrr*(*n*) is a concave function of *n*. We show this in Figure 2. Eq. (20) then immediately follows from Jensen’s inequality. In the high-risk exclusion strategy, the relative risk reduction is neither concave nor convex.

### 5.3 Derivation of the mean risk reduction for a random number of births

Denote by *N* the random variable of the number of births. Our goal is to find

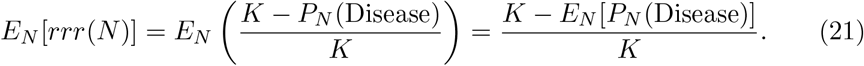

Thus, we need to find *E*_*N*_ [*P*_*N*_ (Disease)].

Denote the probability mass function of *N* as *P* (*N* = *n*). For a given number of births *N*, Eq. (10) provides the risk of the selected embryo,

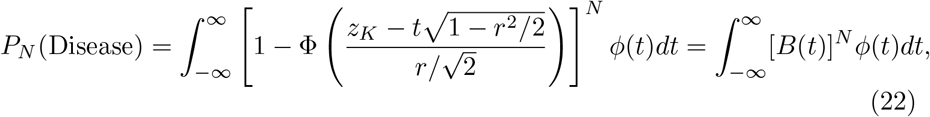

where we defined 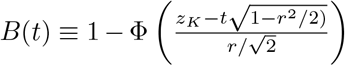, as in Figure 2. The mean risk reduction is

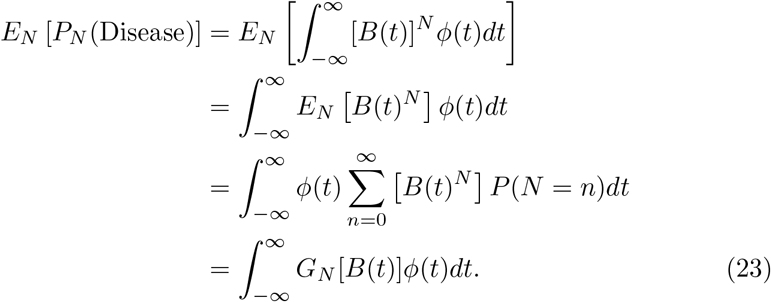

In the last step, we used the probability generating function (PGF) 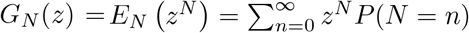.

**Figure S2.** Concavity of the relative risk reduction. Denote by *P*_*n*_(Disease) the probability that the selected embryo (out of *n*) is affected, as computed in Eq. (10) under the lowest risk prioritization strategy. The relative risk reduction is defined as 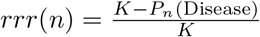. To show that *rrr*(*n*) is concave in *n*, it is sufficient to show that *P*_*n*_(Disease) is convex. Let 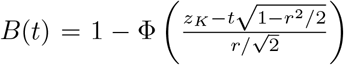. Note 0 < *B*(*t*) < 1. Following Eq. (10), the probability of disease given *n* embryos is 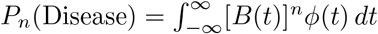. We demonstrate that *P*_*n*_(Disease) is convex in two ways. *Approach 1: Second derivative test* (treating *n* as continuous)

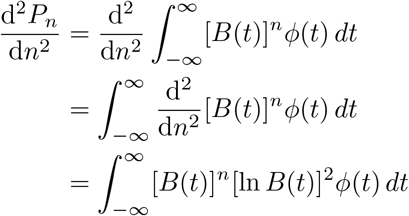

Since [*B*(*t*)]^*n*^ > 0, [ln *B*(*t*)]^2^ ≥ 0, and *ϕ*(*t*) > 0, the integrand is non-negative. Thus, 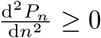, implying that *P*_n_ (Disease) is convex. *Approach 2: Definition of convexity* Let *n* = (1 − *α*)*n*_1_ + *αn*_2_ for *α* ∈ [0, 1].

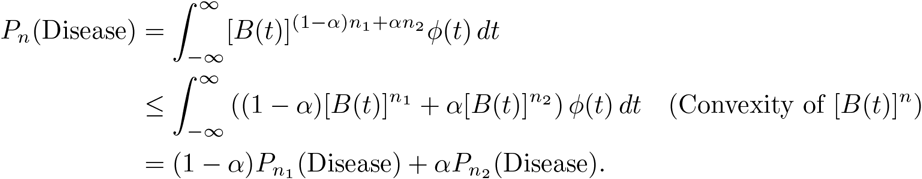

This directly shows *P*_*n*_(Disease) is convex. Both approaches confirm the convexity of *P*_*n*_ and thus the concavity of the risk reduction.

Therefore, Eq. (23) provides a recipe for computing the probability that the selected embryo is affected. For any given distribution of the number of births per IVF cycle (*N*), compute the PGF *G*_*N*_ (*z*), and then solve Eq. (23) numerically.

However, a problem with Eq. (23) is that it also includes the case *N* = 0. But in reality, if there are no births, the risk reduction is not properly defined. In fact, Eq. (10) is nonsensical for *N* = 0. Therefore, in the following, we condition on *N* > 0, implying that IVF cycles are continuously repeated until at least one child is born.

Given the original distribution of *N, P* (*N* = *n*), the probability mass function of the *conditional* number of births is

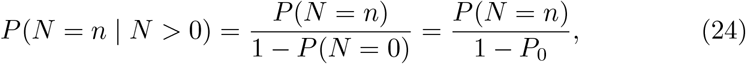

where we defined *P*_0_ *≡ P* (*N* = 0). Further, the PGF becomes

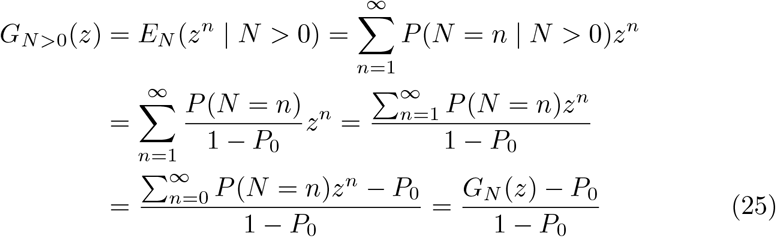

Going back to Eq. (23), we substitute Eq. (25).

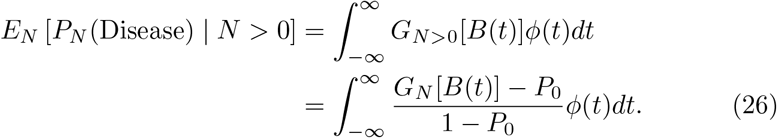

We have thus obtained an integral expression for the risk of the selected embryo, and thus, via Eq. (21), to the mean relative risk reduction *E*_*N*_ [*rrr*(*N*) | *N* > 0]. Eq. (26) depends on *G*_*N*_ (*z*), the PGF of the distribution of *N*. The next step is to consider specific distributions and obtain *E*_*N*_ [*rrr*(*N*) | *N* > 0] for each case.

### 5.4 Case 1: A fixed number of embryos and a binomial number of births

Suppose that the number of (euploid) embryos *n*_0_ is known. For example, this is the case for IVF patients in a given IVF cycle. We then assume that each transferred embryo has an identical probability *p* to be born. Thus, the number of births is binomial, *N* ∼ Bin(*n*_0_, *p*). Note that we do not condition on *N* > 0 (i.e., that at least one embryo was born), since this conditioning is accounted for in Eq. (26).

To use Eq. (26), we substitute the PGF of a binomial random variable,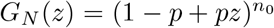. Further,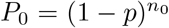. Thus, for a fixed number of embryos and a binomial number of births,

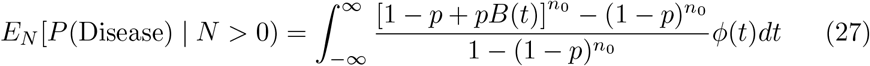

We solve Eq. (27) numerically using R’s `integrate`.

### 5.5 Case 2: A Poisson number of embryos and a binomial number of births

The first case assumed that the number of embryos is fixed, which corresponds to the setting of patients with a specific number of euploid embryos. To obtain the mean risk reduction across the population, it is necessary to take into account the randomness in the number of (euploid) embryos.

In the following, we assume a model whereby the number of euploid embryos has a Poisson distribution with mean *λ*, and then, every transferred embryo is born with probability *p*. We start with the following lemma.

#### Lemma: Poisson followed by binomial

Let

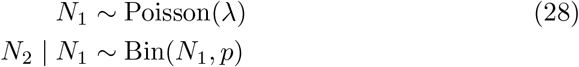

Then *N*_2_ ∼ Poisson(*λp*).

**Proof:** The PGF of a Poisson(*λ*) random variable is *G*_1_(*z*) = *e*^*λ*(*z*−1)^. The PGF of a binomial variable Bin(*N*_1_, *p*) is 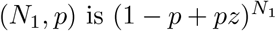. Thus,

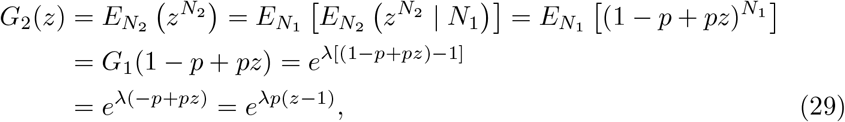

which is the PGF of a Poisson(*λp*) distribution. ¦

To compute the probability that the selected embryo will be affected, we can substitute Eq. (29) (i.e., *G*_*N*_ (*z*) = *e*^*λp*(*z*−1)^ into Eq. (26). Further, *P*_0_ = *P* (*N* = 0) = *e*^−*λp*^. Thus,

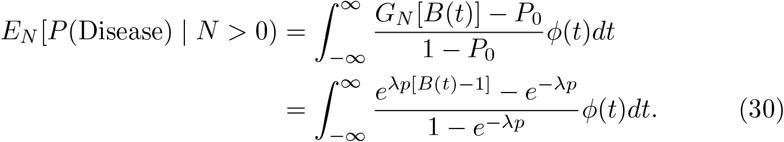

Using a similar approach, it is possible to derive the risk reduction for additional distributions for the number of embryos, such as the negative binomial. However, for the sake of simplicity, we limited the implementation of *PEStimate* only to the Poisson distribution.

### 5.6 A numerical comparison of the three models

We provided above three models for the IVF process:

1. *A fixed number of births*. This model has a single parameter: the number of births *N*. The risk reduction is based on Eq. (22).
2. *A fixed number of embryos and a binomial number of births*. This model has two parameters: the (fixed) number of embryos *n*_0_ and the birth rate *p*. The risk reduction is based on Eq. (27).
3. *A Poisson number of embryos and a binomial number of births*. This model has two parameters: the mean number of embryos *λ* and the birth rate *p*. The risk reduction is based on Eq. (30).

Figure 1 of the main text provides a numerical comparison of the risk reductions predicted by the three models. To allow a fair comparison, the mean number of births must be the same across all models. Due to conditioning on *N* > 0, the mean number of births in the binomial model i s 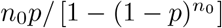. The mean number of births in the Poisson model is *λp/*(1 − *e*^−*λp*^). Thus, we require 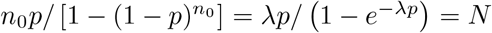. To achieve this, we vary *N* in increments of 1 in the range [1,10]. We then set *p* = 0.4 and vary *n*_0_ or *λ* to achieve equality of the means (accepting the fact that *n*_0_ may be fractional). We further set *r*^2^ = 0.1 and *K* = 0.01.

